# Longitudinal changes in epigenetic clocks predict survival in the InCHIANTI cohort

**DOI:** 10.1101/2024.09.13.24313620

**Authors:** Pei-Lun Kuo, Ann Zenobia Moore, Toshiko Tanaka, Daniel W Belsky, Ake Tzu-Hui Lu, Steve Horvath, Stefania Bandinelli, Luigi Ferrucci

**Affiliations:** National Institute on Aging, National Institutes of Health, Baltimore, MD, USA 21224; Robert N Butler Columbia Aging Center and Department of Epidemiology, Mailman School of Public Health, Columbia University; Altos Labs, San Diego, 5510 Morehouse Dr, San Diego, CA 92121; Dept. of Human Genetics, David Geffen School of Medicine, University of California Los Angeles, Los Angeles, CA, USA 90095; Geriatric Unit, Local Health Unit Tuscany Centre, Firenze, Tuscany 50125, Italy

**Keywords:** Epigenetic clocks, longevity, healthy aging

## Abstract

**Importance:** Cross-sectional assessment of epigenetic clocks provides information on the pace of aging. Whether longitudinal acceleration or deceleration of epigenetic clocks over time provides additional mortality prediction is unknown.

**Objective:** To test the independent associations of baseline levels and longitudinal changes in epigenetic clocks with mortality

**Design:** Longitudinal study

**Setting:** InCHIANTI, a population-based study of community dwelling individuals in Tuscany, Italy.

**Participants:** 699 InCHIANTI study participants aged 21-95 years at baseline with longitudinal measurements of DNA methylation.

**Exposure:** Baseline levels and longitudinal changes in seven epigenetic clocks, including two first-generation clocks developed using chronological age for reference (Hannum Clock, Horvath Clock), three second-generation clocks developed using time-to-death for references (DNAmPhenoAge, DNAmGrimAge, DNAmGrimAge Version 2), and two third-generation clocks developed using longitudinal rate of change of multiple phenotypes for reference (DunedinPOAm_38, DunedinPACE).

**Main Outcomes and Measures:** Mortality was the primary outcome. Cox regression was used to estimate independent associations of baseline and longitudinal changes in epigenetic clocks with mortality.

**Results:** Adjusting for age, sex, study sites, and epigenetic clock at the baseline, longitudinal changes of the following epigenetic clocks were associated with mortality: Hannum clock (aHR = 1.14, 95% CI:[1.03, 1.26]), DNAmPhenoAge (aHR = 1.23, 95% CI: [1.10,1.37]), DNAmGrimAge (aHR = 1.13, 95% CI: [1.02,1.26]), DNAmGrimAge Version 2 (aHR = 1.18, 95% CI:[1.06,1.31]), and DunedinPOAm_38 (aHR = 1.15, 95%CI: [1.01,1.30]).

**Conclusions and Relevance:** Our findings confirm that epigenetic clocks capture a dimension of health that is predictive of mortality and add the notion that time changes of epigenetic age reflect changes in health that additionally and independently contribute to mortality prediction. Future studies should test whether interventions that slow down the rate of epigenetic aging are associated with longer healthspan and lifespan.

**Key Points:** Question: Is the rate of change in epigenetic clock associated with differential mortality?

Findings: In 699 adults with followed for up to 24 years, faster longitudinal changes in epigenetic clocks (Hannum clock, DNAmPhenoAge, DNAmGrimAge, DNAmGrimAge version 2, DunedinPOAm_38, DunedinPACE) were significantly associated with higher mortality, independent of baseline epigenetic age and other confounders.

Meaning: Independent of chronological age, epigenetic clock and change over time of epigenetic clock independently predicted the risk of death. Interventions that slow down the pace of epigenetic aging may enhance healthy longevity.

## Introduction

With continuing progress in living conditions and public health, the population pyramid is shifting towards a top-heavy shape ^1,2^. However, not all the increases in longevity occurs in healthy life, and much is accounted for by people with chronic diseases living longer ^1–3^. A persistent and almost exclusive focus of medicine on disease treatments after their clinical emergence will not change this trend, and will eventually lead to a dramatic expansion of the percentage of the population affected by chronic morbidity and multimorbidity ^3^. The geroscience paradigm may provide an alternative solution to this gloomy scenario. The geroscience poised that the biology of aging is the root cause of chronic diseases and impairments that emerge in late ^4–6^. Thus, slowing down the pace of biological aging may prevent or delay the onset of diseases and disabilities and prolong the healthspan, possibly expanding the proportion of life that people live in good health ^4,5,7,8^. A first step in accomplishing this goal is to develop metrics of biological aging that can identify faster aging individuals and track the effect of interventions aimed at slowing down the aging process ^4,7,9^.

Over the past years, several proxy biomarkers of biological aging were developed and validated, with the most advanced using data from DNA methylation data ^10^. In general, these epigenetic markers of aging (later referred as “epigenetic clocks”) predict multiple adverse health outcomes, including mortality independent of chronological age ^11^. Early in the 2010s, the Hannum clock and Horvath clock were developed combining information on percent methylation in selected DNA sites to obtain a score that best correlated with chronological age ^12,13^. The second-generation clocks such as DNAmPhenoAge, DNAmGrimAge, DNAmGrimAge version 2 were developed by using mortality and other blood biomarkers for reference ^14–16^. The DunedinPOAm_38 and DunedinPACE were developed using the pace biological aging estimated from a combination of longitudinal trends of multiple phenotypes in a birth cohort for reference ^17,18^. However, whether longitudinal changes in these phenotypes provide additional information on health outcome prediction over and one single measure has not been demonstrated ^7,19^. Based on cross-sectional studies, we cannot definitively exclude that deviations of DNA methylation age from chronological age are determined early in life and are not modulated by behavioral, environmental exposures or changes in health status ^7,20,21^. In addition, if biological aging clocks are to be used to track the effectiveness of intervention over time, it is important to demonstrate that deviations of epigenetic clock trajectories reflect meaningful changes in health status ^22^. In this study, we use longitudinal data collected in the InCHIANTI study to test the hypothesis that longitudinal changes in epigenetic clock predict mortality over and beyond the same epigenetic clock assessed at one point in time.

## Methods

### Study Design and Participants

The InCHIANTI study participants were recruited from two Chianti regions of Tuscany, Italy, (1998 – 2000). Details about this study have been described previously ^23^. For the present analysis, we included 699 participants with DNA methylation measurements measured at two follow-up or more timepoints (in the 1998, 2007, and 2013 study visits). InCHIANTI was approved by the Italian National Institute of Research and Care of Aging Institutional Review Board and study participants provided informed consents. This study adheres to the Strengthening the Reporting of Observational Studies in Epidemiology (STROBE) reporting guideline ^24^.

### Epigenetic Markers of Aging ("Epigenetic Clocks”)

DNA methylation was measured using the Illumina Infinium HumanMethylation450 BeadChip ^25^. Data quality control included background correction, multi-dimensional scaling, and checking that self-reported sex was consistent the estimated copy number of sex chromosomes ^26^. R packages "minfi" and "SeSAMe" were used for quality control and to generate methylation beta values, which represent levels of methylation and range between 0 (completely unmethylated) and 1 (completely methylated) ^26–28^. A total of 1721 samples from 699 participants (376 participants measured at two timepoints, 323 participants measured at three timepoints) were used in the analysis.

We used DNA methylation data to calculate seven epigenetic clocks. The first-generation clocks (Hannum clock, Horvath clock) and the second-generation clocks (DNAmPhenoAge and two versions of DNAmGrimAge) were calculated through web-based calculator (https://horvath.genetics.ucla.edu/html/dnamage/). Because these five epigenetic clocks were highly associated with chronological age, in cross-sectional studies, the age-adjusted values of them were often called “epigenetic age acceleration” (despite not being measured longitudinally) and used as the “baseline epigenetic clocks”. The third-generation epigenetic clocks (DunedinPOAm_38 and DunedinPACE), which are developed to estimate the pace of biological aging and thus different from other clocks on the interpretation and scale, were calculated using DunedinPOAm_38, and DunedinPACE R packages (https://github.com/danbelsky/DunedinPoAm38 and https://github.com/danbelsky/DunedinPACE) ^17,18^.

Annual rate of changes in epigenetic clocks for each individual were estimated by linear regression analyses. Because different epigenetic clocks were derived differently and came with different scales, all the baseline level and longitudinal changes in epigenetic clocks were standardized to the unit of standard deviation (SD) so to allow a fair comparison.

A recent literature proposed that epigenetic clocks estimated by principal-component analysis (“PC clocks”) have better psychometric properties than the original versions ^22^. Therefore, we conducted the analysis using the PC versions of the clocks ^22,29^. Additionally, since DNAmGrimAge version 2 contained several protein proxies, we also reported the associations between these protein proxies and mortality.

### Other Independent Variables

Self-reported sex, chronological age, and sites of residency were collected during the study visit. A composite score of cardiovascular health was calculated using information on the life simple 7 (LS7), namely smoking, physical activity, body mass index, total cholesterol, blood pressure, fasting plasma glucose and adherence to a Mediterranean-style diet ^30^. The score ranged from 0 to 12, with higher numbers reflecting better cardiovascular health ^30^.

### Outcome Measure

Vital status was ascertained by linkage with the Registry office of the municipality of residency from the day after the baseline interview up to the end of January 2023).

### Statistical Analysis

We used Cox regression to examine the relationships between standardized values of epigenetic clocks and mortality across three models: "Baseline epigenetic clock only", "Change over time of epigenetic clock only", and "Both baseline and change over time of epigenetic clock". All models were adjusted for baseline chronological age, sex, and study site and subsequently further adjusted for the Life Simple Seven score. Results are shown by forest plots and reported C-statistics for all three models and nested models are compared by likelihood ratio tests. Hazard ratios fail to provide a direct measure of how well biomarkers can classify risk, and C-statistics may not be sufficiently sensitive to detect meaningful improvements, we also calculated the integrated discrimination index (IDI) and net reclassification index (NRI), which have been proposed as more sensitive alternatives to evaluate improvement in risk prediction ^31–33^. As over 40% of participants were administratively censored at the end of follow-up, we used the R package "survIDINRI" by Uno et al. to calculate IDI and NRI at 20 years with 1,000 bootstrap sampling. To estimate the age-specific rate-of change in epigenetic clocks, we used linear mixed model with random intercept and random slope, compared models using polynomial curves with different degrees of freedom, and chose the model with the lowest Akaike Information Criterion. We conducted our analysis and plotting in SAS 9.4, and R version 3.6.2, using the "survival" package for survival analysis. All point estimates were reported with 95% confidence interval.

## Results

The study population included 699 participants, of whom 308 (44%) were male. Of this initial cohort, 396 participants died over 24 years follow up (mortality rate = 29.14 deaths / 1,000 person-years). Median (range) of follow-up time was 21.5 (IQR = 15.6 – 23.4) years. The mean baseline chronological age of the participants was 63 (SD=16) years old. At baseline, the lowest mean value for epigenetic aging markers was 55.9 years (SD=17.4) for DNAmPhenoAge while the highest mean was 67.2 years (SD=12.9) for DNAmGrimAge version 2. The mean values for DunedinPOAm_38 and DunedinPACE were 1.02 (SD=0.09) and 1.06 (SD=0.12). Across the other epigenetic markers of aging the smallest mean annual rate of change was observed for DNAmGrimAge version 2, 0.66 (SD=0.29), while DNAmPhenoAge had the largest value, 0.96 (SD=0.50) (Table 1). The correlations between baseline epigenetic markers of aging and annual rates of change in epigenetic markers of aging ranged from -0.20 to -0.49 (Supplemental Figure 1-3).

**Table 1.**
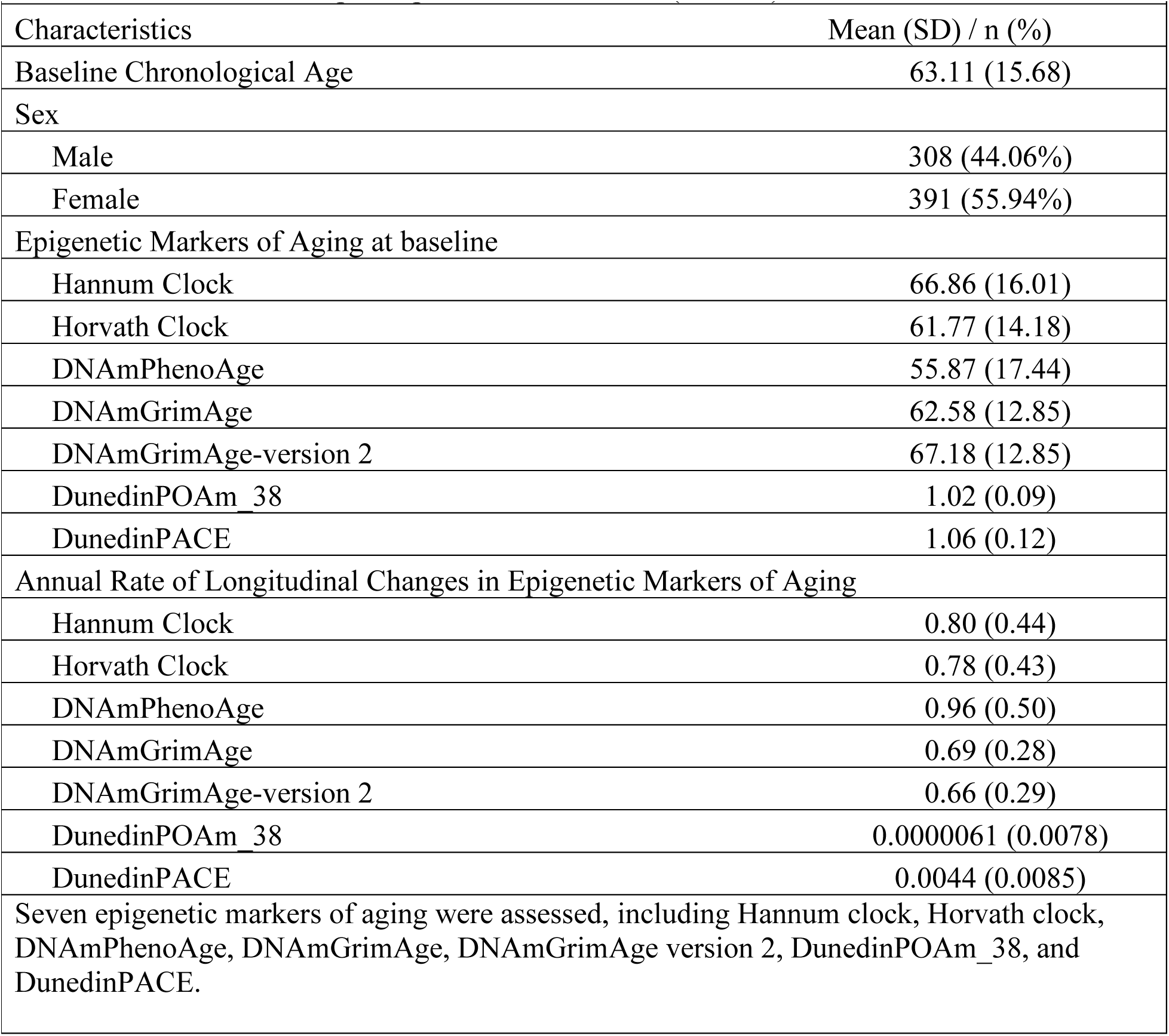
Characteristics of participants in InCHIANTI (n = 699)

Figure 1 shows “spaghetti” plots illustrating longitudinal changes in epigenetic clocks for 699 participants with repeated-measures data. The consistency of linear change over time within individuals is striking, with only a few individuals showing strong deviations. Interestingly, some clocks appear to tick slower/faster than others. The slowest ticking rate was observed for DNAmGrimAge version 2. The fastest ticking rate was observed for DNAmPhenoAge.

**Figure 1.**
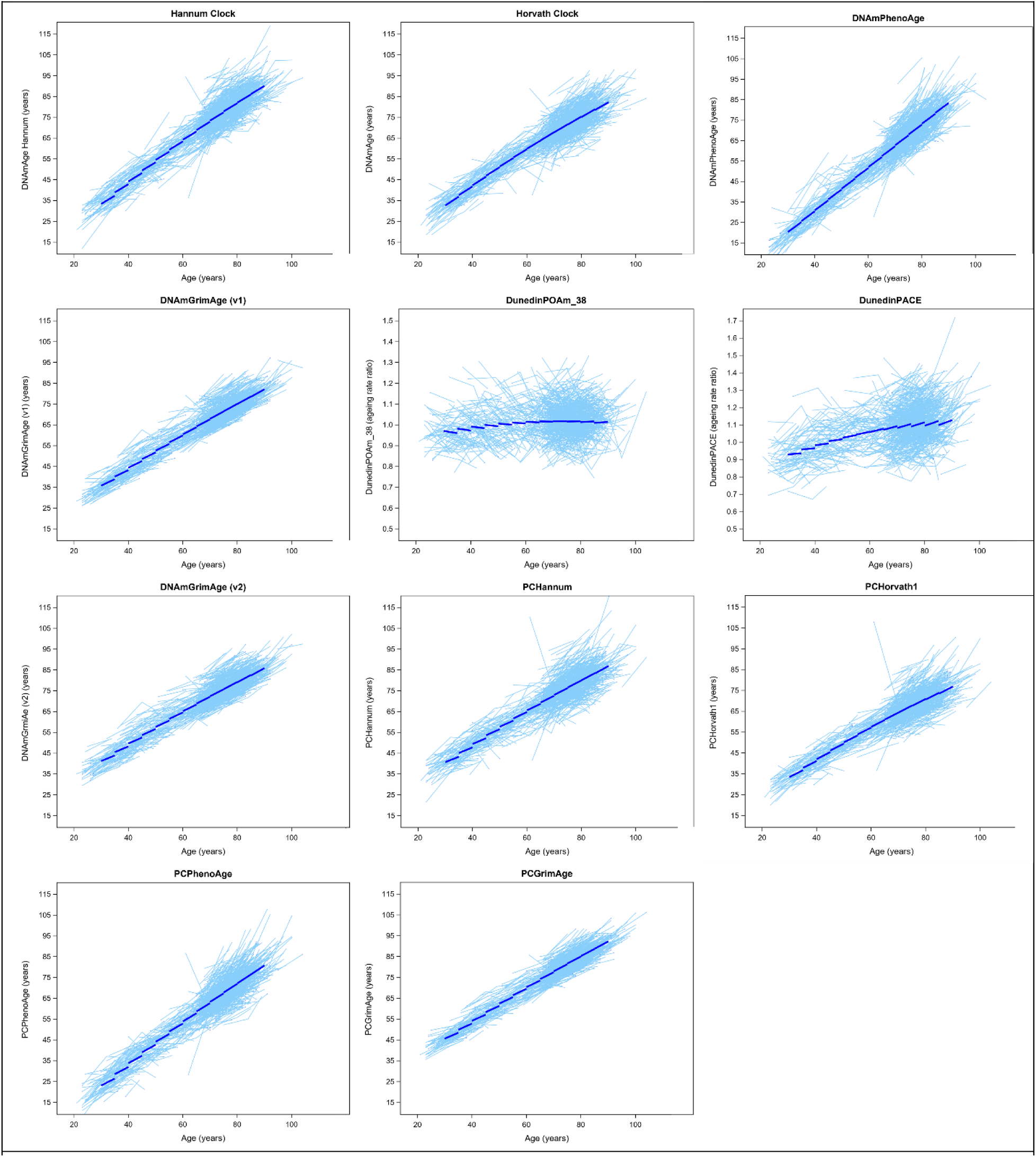
Longitudinal trajectories of epigenetic clocks. For the Figure 1, each light blue line represents the longitudinal repeated measurements for an individual, and the dark blue line represents the fitted trajectories across all ages.

DunedinPOAm_38 and DunedinPACE already express pace of epigenetic aging, and therefore longitudinal trajectories over time should be interpreted as age-accelerations or deceleration.

Interestingly, DunedinPOAm_38 trajectories showed relative stable pace of epigenetic aging across different age groups while the DunedinPACE trajectories showed DundeinPACE increased over time, which is consistent with previous reports ^18,34^. In addition, the rate of increase in DunedinPACE accelerated as participants got older (Supplemental Table 2). Estimated age-specific annual rates of change in epigenetic age for different epigenetic clocks are shown in Supplemental Table 2.

The forest plots in Figures 2 and 3 provide visual representations of the relationship between epigenetic clocks and mortality. In the "Baseline only" model, all epigenetic markers of aging except for Horvath clock were associated with mortality (aHR [95% CI]: 1.12 [1.01 – 1.23] for Hannum clock, 1.06 [0.96 – 1.17] for Horvath clock, 1.22 [1.10 – 1.35] for DNAmPhenoAge, 1.38 [1.23 – 1.55] for DNAmGrimAge, 1.38 [1.23 – 1.55] for DNAmGrimAge version 2, 1.19 [1.06 – 1.33] for DundedinPOAm_38, and 1.23 [1.10 – 1.38] for DunedinPACE). In the "Slope only" model, only the longitudinal changes in Hannum clock and DNAmPhenoAge were associated with mortality (aHR [95% CI]: 1.10 [1.00 – 1.24] for Hannum clock, 0.99 [0.89 –1.09] for Horvath clock, 1.12 [1.01 – 1.24] for DNAmPhenoAge, 1.01 [0.91 – 1.13] for DNAmGrimAge, 1.08 [0.97 – 1.21] for DNAmGrimAge version 2, 1.01 [0.91 – 1.13] for DundedinPOAm_38, and 1.02 [0.92 – 1.14] for DunedinPACE). When both baseline and longitudinal changes were included in the model (Figure 3), the associations between baseline measures and mortality were generally stronger than the estimates from the "Baseline only" model, and the associations between longitudinal changes and mortality were generally stronger than the estimates from the "Slope only" model (Figure 2). Results remained substantially unchanged after additional adjustment of Life Simple Seven (Supplemental Figure 4). When we repeated the analysis using PC clocks (the epigenetic clocks calculated adopting technical noises reduction algorithms), the associations between changes in epigenetic clocks and mortality using PC clocks were very similar to those obtained using the original clocks, except for an observable improvement in Horvath Clock (Supplemental Figure 5). Interestingly, we explored whether the epigenetic estimators of circulating proteins included in DNAmGrimAge version 2 were associated with mortality and found that longitudinal changes in methylation-based estimation GDF-15, Cystatin-C, TIMP-1, and logCRP were associated with mortality (Supplemental Table 3).

**Figure 2.**
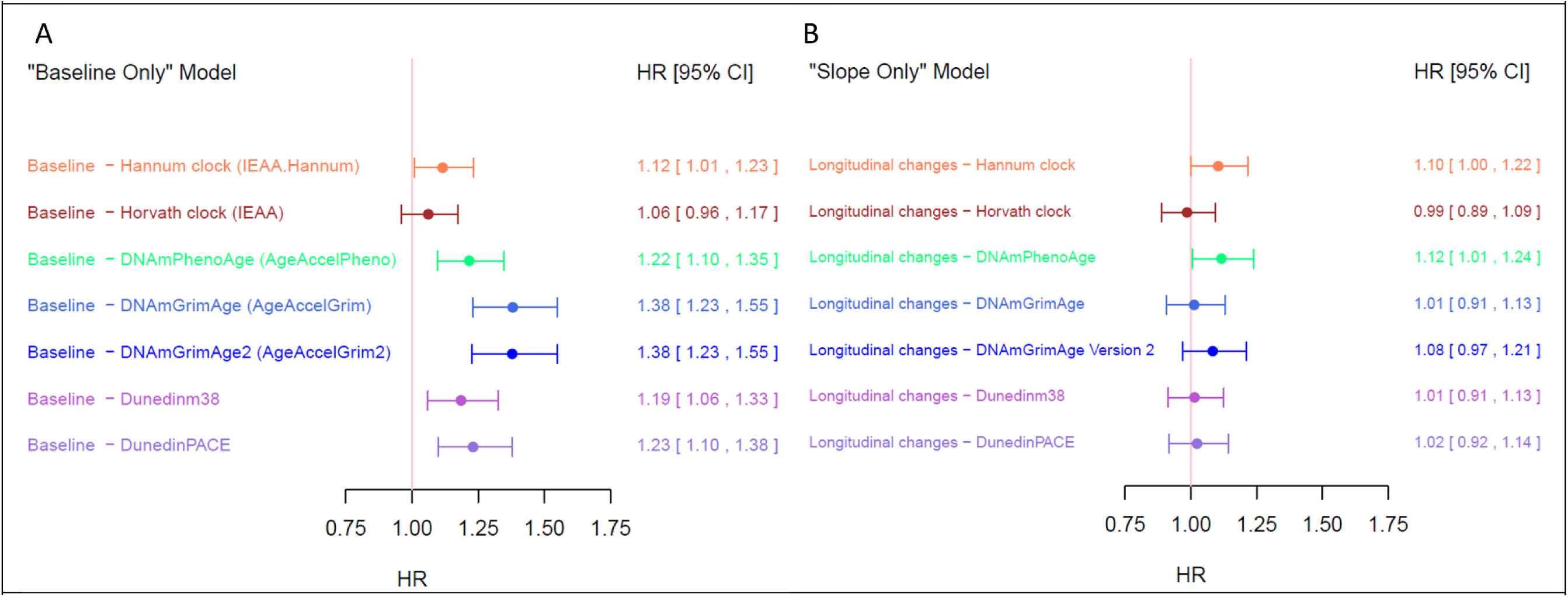
Forest Plot for the Adjusted Hazard Ratios of Mortality. For the Figure 2A, the hazard ratios are derived from the model including baseline epigenetic markers of aging, chronological age, sex, and study site. For the Figure 2B, the hazard ratios are derived from the model including annual rate of longitudinal changes in epigenetic markers of aging, chronological age, sex, and study site. To facilitate the interpretation and comparison, all the coefficients are standardized.

**Figure 3.**
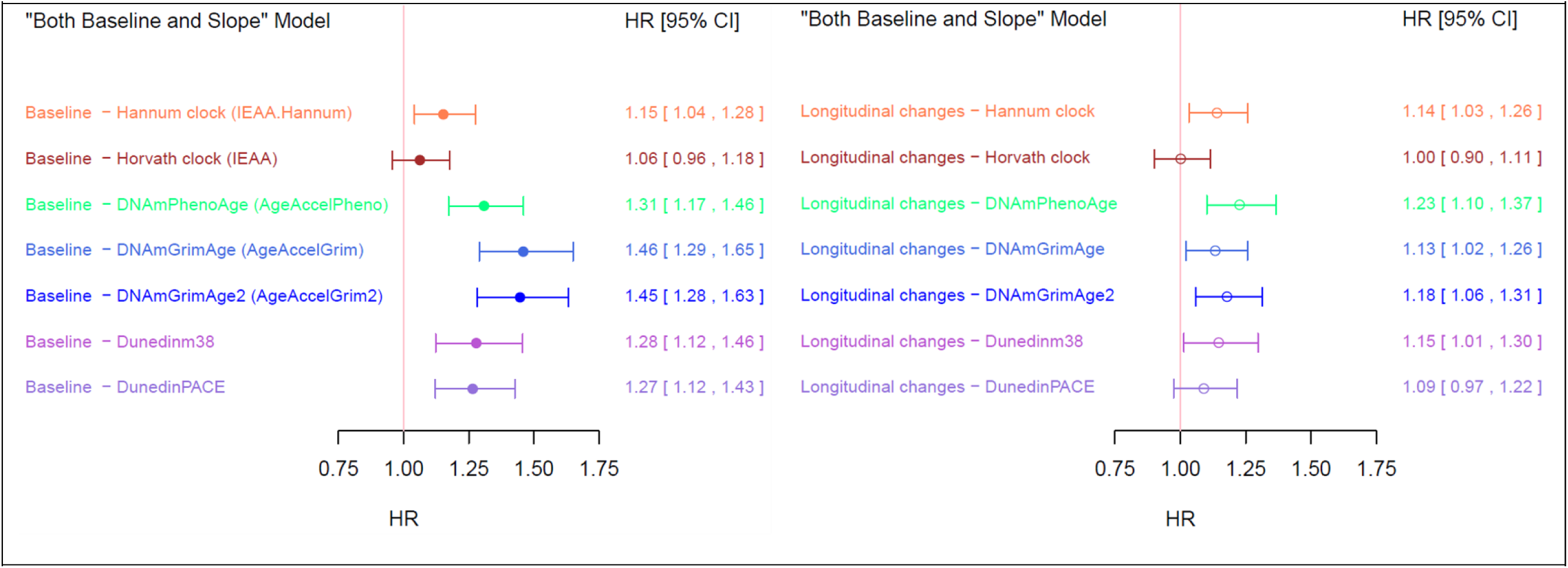
Forest Plot for the Adjusted Hazard Ratios of Mortality when both Baseline and Longitudinal Changes are included in the model. For the Figure 3, the hazard ratios are derived from the model including baseline epigenetic markers of aging, annual rate of longitudinal changes in epigenetic markers of aging, chronological age, sex, and study site. To facilitate the interpretation and comparison, all the coefficients are standardized.

The performance of the different models for mortality prediction was assessed using a variety of measures, including c-statistics, integrated discrimination improvement index (IDI), and net reclassification index (NRI). The results are presented in Supplemental Table 1 and Table 2. The models that included both baseline and longitudinal change for DNAmGrimAge version 2, DNAmGrimAge, DNAmPhenoAge, and DunedinPACE had the highest c-statistics (Concordance index: 0.808 for DNAmGrimAge version 2, 0.806 for DNAmGrimAge, 0.801 for DNAmPhenoAge, 0.800 for DunedinPACE). The integrated discrimination improvement index and net reclassification index also showed that the integration of baseline value and longitudinal changes in epigenetic clocks demonstrated improvement in mortality prediction performance (IDI: 0.017 [0.005 – 0.035] for DNAmPhenoAge, 0.027 [0.010 – 0.045] for DNAmGrimAge; NRI: 0.251 [0.084 – 0.320] for DNAmPhenoAge, 0.236 [0.053 – 0.316] for DNAmGrimAge).

**Table 2.** Integrated Discrimination Improvement and Net Reclassification Performance of Models including Epigenetic Markers of Aging.

**Title:** Longitudinal changes in epigenetic clocks predict survival in the InCHIANTI cohort
|  |  |  |
| --- | --- | --- |
| Epigenetic Markers of Aging | IDI | NRI |
| Hannum Clock | **0.010 [ 0.000, 0.026]** | 0.186 [ -0.161, 0.300] |
| Horvath Clock | 0.001 [ -0.001, 0.009] | 0.152 [ -0.204, 0.237] |
| DNAmPhenoAge | **0.017 [ 0.005, 0.035]** | **0.251 [ 0.084, 0.320]** |
| DNAmGrimAge | **0.027 [ 0.010, 0.045]** | **0.236 [ 0.053, 0.316]** |
| DNAmGrimAge version 2 | **0.029 [ 0.013, 0.051]** | **0.198 [ 0.003, 0.291]** |
| DunedinPOAm_38 | 0.009 [ -0.001, 0.024] | 0.126 [ -0.087, 0.222] |
| DunedinPACE | **0.010 [ 0.001, 0.027]** | 0.035 [ -0.127, 0.158] |
| The IDI and NRI are used to assess the additional prediction values of models including epigenetic models of aging. Specifically, it compares the following two models for each epigenetic markers of aging: (i) model including chronological age, sex, and study site, and (ii) model including “baseline epigenetic markers of aging”, “annual rate of longitudinal changes in epigenetic markers of aging”, chronological age, sex, and study site.<br>IDI = Integrated Discrimination Improvement Index; NRI = net reclassification index |  |  |

## Discussion

We found that changes in most epigenetic clocks are predict differential mortality, independent of chronological age, baseline epigenetic clock, and sex. Second-generation clocks trained using mortality for reference (DNAmPhenoAge, DNAmGrimAge, DNAmGrimAge version 2) or third-generation clocks trained against longitudinal phenotypic changes (DunedinPOAm_38, DunedinPACE) performed better for mortality prediction than first-generation clocks, as evidenced by higher concordance indices and integrated discrimination improvement indices.

Specifically, second-generation clocks (DNAmPhenoAge, DNAmGrimAge, DNAmGrimAge version 2) had the highest net reclassification indexes in our study cohort. To our knowledge, this is the first study to examine the association between longitudinal changes in the epigenetic most used in the literature and mortality.

The association between baseline epigenetic clock and mortality was previously reported by many studies. Higgins-Chen and colleagues showed that DNAmPhenoAge and DNAmGrimAge had a stronger association with mortality than the Hannum clock and Horvath clock, even after improving their reliability using novel computational techniques ^22^. Similarly, Belsky and colleagues found that DunedinPOAm_38 and DunedinPACE were more strongly associated with mortality than Hannum clock and Horvath clock, consistent with our results ^17,18^. By leveraging longitudinal data, we were able to include both baseline and longitudinal measurements of epigenetic clocks in our analyses and demonstrated that the mortality prediction was substantially higher when both baseline and longitudinal estimates of epigenetic aging were considered jointly. A puzzling result was that different epigenetic clocks showed different longitudinal slopes of epigenetic aging, all of which were lower than one epigenetic year per year of chronological age. The reason for this finding is unclear. In the future, epigenetic clocks estimated using repeated measures of methylation and correlating them with change in age- phenotypes may shed light on this result.

Longitudinal changes in epigenetic clock without adjusting for baseline value were associated with mortality only for the Hannum clock and DNAmPhenoAge but not for the other clocks. These findings cannot be directly compared with the literature because the literature exploring longitudinal changes in epigenetic markers is scant ^34,35^. We propose that epigenetic age is not a straight trajectory over time but show deviations from this overall trajectory in response to meaningful changes in health, and that is why considering such deviation improves our mortality prediction. A definitive demonstration of this hypothesis would require larger study, with longer follow-up and methylation data available in multiple visits so that fluctuations in epigenetic age can be correlated to exposures and health events.

In our analysis, second-generation (DNAmPhenoAge, DNAmGrimAge, DNAmGrimAge version 2) and third-generation (DunedinPOAm_38, DunedinPACE) clocks were superior to the first-generation clocks (Hannum clock, Horvath clock) in mortality prediction. This was not unexpected since part of second-generation clocks were optimized to predict mortality, and third-generation clocks were built upon the longitudinal changes of many clinically important phenotypes. In contrast, first generation clocks were trained on chronologic age, which obscure deviation from the biological age that may convey important information on mortality. Second, DNAmPhenoAge and DNAmGrimAge appeared to have better net reclassification ability than DunedinPOAm_38 and DunedinPACE. One plausible reason is that the optimization of DunedinPOAm_38 and DunedinPACE did not involve mortality information at all. The other plausible reason is that DNAmPhenoAge and DNAmGrimAge were trained on adults across a wider age range than DunedinPOAm_38 and DunedinPACE, which were trained on younger adults at 26-38 and 26-45, respectively, and their predictive validity may not fully extend to older age ^17,18^.

Our study supports the notion that longitudinal trajectories of epigenetic clocks can be reliably used in observational studies to detect important deviations of health due to intrinsic or extrinsic factors that ultimately are reflected by differential mortality. On the other hand, our findings reinforce idea that epigenetic clocks can be used to test the effectiveness of interventions aimed at slowing down biological aging and prevent its deleterious effects ^6^. While our study focused on mortality, it will be important to expand these observations to different outcomes that are theoretically associated with accelerated aging, such as multimorbidity and disability ^7,8^. Future research should also investigate whether interventions that slow the changes in epigenetic markers of aging can improve health span and life expectancy. Ultimately, it will be important to understand what biological mechanism drives the changes in the trajectories of epigenetic clocks and the phenotypes correlate with these changes.

There are several strengths and limitations in our study. All participants were of European ancestry. Most epigenetic clocks used in research have been developed from European ancestry data. Hence, one important goal of future research is to develop epigenetic clocks from a highly diverse populations and verify that their predictive validity is similar across race and sex. The lack of overlapping CpG sites among these epigenetic clocks and the poor correlation among them reported in the literature suggests that they may convey much more information than we can currently summarize. Thus, larger studies with the use of artificial intelligence to unveil underlying epigenetic networks may be extremely helpful to refine these tools and make them more robust to translation in clinical applications. Nevertheless, our results provide the first evidence that longitudinal changes in epigenetic clock values reflect meaningful changes in health, lay out the background for expanding and translating this line of research.

## Data Availability

“The InCHIANTI database is available to any researcher who presents a detailed and scientifically sound project. Data available through InCHIANTI cover a range of factors that influence walking ability that are classified into the six subsystems listed above. Measures of the integrity and functioning of each of these subsystems were identified and administered to all participants.”
Link to access: “https://www.nia.nih.gov/inchianti-study#access”

https://www.nia.nih.gov/inchianti-study#access

