## Supplements for "Longitudinal changes in epigenetic clocks predict survival in the InCHIANTI cohort"

| Supplemental Table 1. C-statistics of models for epigenetic markers of aging | | |
|---|---|---|
| Survival Models (*) | | |
| Hannum Clock | Concordance index | P-value (**) |
| Baseline only | 0.7958 | 0.010 |
| Slope only | 0.7944 | 0.007 |
| Both Baseline and Slope | 0.7983 | Ref |
| Horvath Clock | | |
| Baseline only | 0.7940 | 0.975 |
| Slope only | 0.7926 | 0.251 |
| Both Baseline and Slope | 0.7939 | Ref |
| DNAmPhenoAge | | |
| Baseline only | 0.7978 | <0.001 |
| Slope only | 0.7946 | <0.001 |
| Both Baseline and Slope | 0.8006 | Ref |
| DNAmGrimAge | | |
| Baseline only | 0.8039 | 0.019 |
| Slope only | 0.7928 | <0.001 |
| Both Baseline and Slope | 0.8063 | Ref |
| DNAmGrimAge version 2 | | |
| Baseline only | 0.8039 | 0.003 |
| Slope only | 0.7942 | <0.001 |
| Both Baseline and Slope | 0.8075 | Ref |
| DunedinPOAm_38 | | |
| Baseline only | 0.7961 | 0.029 |
| Slope only | 0.7931 | <0.001 |
| Both Baseline and Slope | 0.7993 | Ref |
| DunedinPACE | | |
| Baseline only | 0.7985 | 0.130 |
| Slope only | 0.7932 | <0.001 |
| Both Baseline and Slope | 0.8004 | Ref |

(*) All the survival models include chronological age, sex, and site. For each epigenetic markers of aging, we fit three models (**“Baseline only”**, **“Slope only”**, and **“Both Baseline and Slope”**). **“Baseline only”** refers to the model including baseline epigenetic markers of aging, chronological age, sex, and study site. **“Slope only”** refers to the model including annual rate of longitudinal changes in epigenetic markers of aging, chronological age, sex, and study site. **“Both Baseline and Slope”** refers to the model including baseline epigenetic markers of aging, annual rate of longitudinal changes in epigenetic markers of aging, chronological age, sex, and study site.
(**) Because the **“Baseline only”** and **“Slope only”** models are nested in **“Both Baseline and Slope”** model, likelihood ratio test is used for model comparison with **“Both Baseline and Slope”** as the reference.

**Title**: Longitudinal changes in epigenetic clocks predict survival in the InCHIANTI cohort

Supplemental Table 2. Estimated Age-specific Annual Rate of Change in Epigenetic Clocks

| | Estimated Annual Rate of Change in Epigenetic Clocks (year of epigenetic age per 1 year increase in chronological age) | | | | |
|---|---|---|---|---|---|
| Age | Hannum | Horvath | DNAmPhenoAge | DNAmGrimAge (v1) | DNAmGrimAge-version 2 |
| 50 | 0.79 [ 0.76, 0.82 ] | 0.82 [ 0.77, 0.86 ] | 0.97 [ 0.93, 1.00 ] | 0.65 [ 0.62, 0.68 ] | 0.59 [ 0.57, 0.62 ] |
| 55 | 0.79 [ 0.76, 0.82 ] | 0.81 [ 0.77, 0.84 ] | 0.97 [ 0.93, 1.00 ] | 0.66 [ 0.63, 0.68 ] | 0.61 [ 0.59, 0.64 ] |
| 60 | 0.79 [ 0.76, 0.82 ] | 0.80 [ 0.77, 0.83 ] | 0.97 [ 0.93, 1.00 ] | 0.67 [ 0.65, 0.69 ] | 0.63 [ 0.61, 0.65 ] |
| 65 | 0.79 [ 0.76, 0.82 ] | 0.79 [ 0.76, 0.82 ] | 0.97 [ 0.93, 1.00 ] | 0.68 [ 0.66, 0.70 ] | 0.65 [ 0.63, 0.66 ] |
| 70 | 0.79 [ 0.76, 0.82 ] | 0.78 [ 0.75, 0.81 ] | 0.97 [ 0.93, 1.00 ] | 0.69 [ 0.67, 0.71 ] | 0.66 [ 0.64, 0.68 ] |
| 75 | 0.79 [ 0.76, 0.82 ] | 0.77 [ 0.74, 0.80 ] | 0.97 [ 0.93, 1.00 ] | 0.70 [ 0.68, 0.72 ] | 0.68 [ 0.66, 0.70 ] |
| 80 | 0.79 [ 0.76, 0.82 ] | 0.76 [ 0.72, 0.79 ] | 0.97 [ 0.93, 1.00 ] | 0.71 [ 0.69, 0.73 ] | 0.70 [ 0.67, 0.72 ] |
| 85 | 0.79 [ 0.76, 0.82 ] | 0.75 [ 0.71, 0.79 ] | 0.97 [ 0.93, 1.00 ] | 0.72 [ 0.69, 0.75 ] | 0.71 [ 0.69, 0.74 ] |
| 90 | 0.79 [ 0.76, 0.82 ] | 0.74 [ 0.69, 0.79 ] | 0.97 [ 0.93, 1.00 ] | 0.73 [ 0.70, 0.76 ] | 0.73 [ 0.70, 0.76 ] |

| | Estimated Annual Rate of Change in Epigenetic Clocks (change in “pace of aging” per 1 year increase in chronological age) | |
|---|---|---|
| Age | DunedinPOAm_38 | DunedinPACE |
| 50 | -0.001210 [ -0.002019, -0.000401 ] | 0.002630 [ 0.001791, 0.003470 ] |
| 55 | -0.000928 [ -0.001618, -0.000239 ] | 0.003056 [ 0.002341, 0.003771 ] |
| 60 | -0.000646 [ -0.001240, -0.000053 ] | 0.003482 [ 0.002867, 0.004098 ] |
| 65 | -0.000365 [ -0.000901, 0.000172 ] | 0.003908 [ 0.003353, 0.004463 ] |
| 70 | -0.000083 [ -0.000612, 0.000446 ] | 0.004334 [ 0.003788, 0.004881 ] |
| 75 | 0.000199 [ -0.000375, 0.000773 ] | 0.004760 [ 0.004168, 0.005352 ] |
| 80 | 0.000481 [ -0.000179, 0.001141 ] | 0.005186 [ 0.004506, 0.005867 ] |
| 85 | 0.000763 [ -0.000011, 0.001537 ] | 0.005612 [ 0.004814, 0.006410 ] |
| 90 | 0.001045 [ 0.000139, 0.001950 ] | 0.006038 [ 0.005104, 0.006972 ] |

**Title**: Longitudinal changes in epigenetic clocks predict survival in the InCHIANTI cohort

| Age | Estimated Annual Rate of Change in Epigenetic Clocks (year of epigenetic age per 1 year increase in chronological age) | | | |
|---|---|---|---|---|
| | PCHannum | PCHorvath1 | PCPhenoAge | PCGrimAge |
| 50 | 0.58 [ 0.53, 0.63 ] | 0.67 [ 0.64, 0.69 ] | 0.74 [ 0.69, 0.78 ] | 0.61 [ 0.59, 0.64 ] |
| 55 | 0.60 [ 0.56, 0.64 ] | 0.67 [ 0.64, 0.69 ] | 0.75 [ 0.71, 0.80 ] | 0.63 [ 0.61, 0.64 ] |
| 60 | 0.62 [ 0.58, 0.66 ] | 0.67 [ 0.64, 0.69 ] | 0.77 [ 0.74, 0.81 ] | 0.64 [ 0.62, 0.65 ] |
| 65 | 0.64 [ 0.60, 0.67 ] | 0.67 [ 0.64, 0.69 ] | 0.79 [ 0.76, 0.82 ] | 0.65 [ 0.64, 0.67 ] |
| 70 | 0.65 [ 0.62, 0.69 ] | 0.67 [ 0.64, 0.69 ] | 0.81 [ 0.78, 0.84 ] | 0.66 [ 0.65, 0.68 ] |
| 75 | 0.67 [ 0.63, 0.71 ] | 0.67 [ 0.64, 0.69 ] | 0.83 [ 0.79, 0.86 ] | 0.68 [ 0.66, 0.69 ] |
| 80 | 0.69 [ 0.65, 0.73 ] | 0.67 [ 0.64, 0.69 ] | 0.84 [ 0.81, 0.88 ] | 0.69 [ 0.67, 0.71 ] |
| 85 | 0.70 [ 0.66, 0.75 ] | 0.67 [ 0.64, 0.69 ] | 0.86 [ 0.82, 0.91 ] | 0.70 [ 0.68, 0.72 ] |
| 90 | 0.72 [ 0.67, 0.78 ] | 0.67 [ 0.64, 0.69 ] | 0.88 [ 0.83, 0.93 ] | 0.71 [ 0.69, 0.74 ] |

**Title**: Longitudinal changes in epigenetic clocks predict survival in the InCHIANTI cohort

Supplemental Table 3. Adjusted Hazard Ratios of Mortality for the protein proxy in GrimAge version 2 when both Baseline and Longitudinal Changes are included

| Variable Name | HR [95% CI] | Variable Name | HR [95% CI] |
|---|---|---|---|
| Baseline - DNAmadm | **1.32 [ 1.05 , 1.65 ]** | Longitudinal Changes - DNAmadm | 1.07 [ 0.95 , 1.21 ] |
| Baseline - DNAmB2M | **1.45 [ 1.06 , 1.98 ]** | Longitudinal Changes - DNAmB2M | 1.06 [ 0.94 , 1.20 ] |
| Baseline - DNAmCystatin_C | **1.64 [ 1.25 , 2.15 ]** | Longitudinal Changes - DNAmCystatin_C | **1.15 [ 1.02 , 1.29 ]** |
| Baseline - DNAmGDF_15 | **1.72 [ 1.39 , 2.13 ]** | Longitudinal Changes - DNAmGDF_15 | **1.09 [ 1.00 , 1.18 ]** |
| Baseline - DNAmleptin | 0.94 [ 0.73 , 1.20 ] | Longitudinal Changes - DNAmleptin | 0.93 [ 0.81 , 1.06 ] |
| Baseline - DNAmlogA1C | 0.92 [ 0.81 , 1.05 ] | Longitudinal Changes - DNAmlogA1C | 1.01 [ 0.90 , 1.13 ] |
| Baseline - DNAmlogCRP | **1.20 [ 1.06 , 1.35 ]** | Longitudinal Changes - DNAmlogCRP | **1.21 [ 1.08 , 1.36 ]** |
| Baseline - DNAmpai_1 | 0.98 [ 0.87 , 1.12 ] | Longitudinal Changes - DNAmpai_1 | 1.02 [ 0.91 , 1.15 ] |
| Baseline - DNAmTIMP_1 | **2.13 [ 1.28 , 3.54 ]** | Longitudinal Changes - DNAmTIMP_1 | **1.17 [ 1.05 , 1.31 ]** |

**Title**: Longitudinal changes in epigenetic clocks predict survival in the InCHIANTI cohort

Supplemental Figure 1. Scatterplot of baseline epigenetic markers of aging and annual rate of longitudinal changes in epigenetic markers of aging

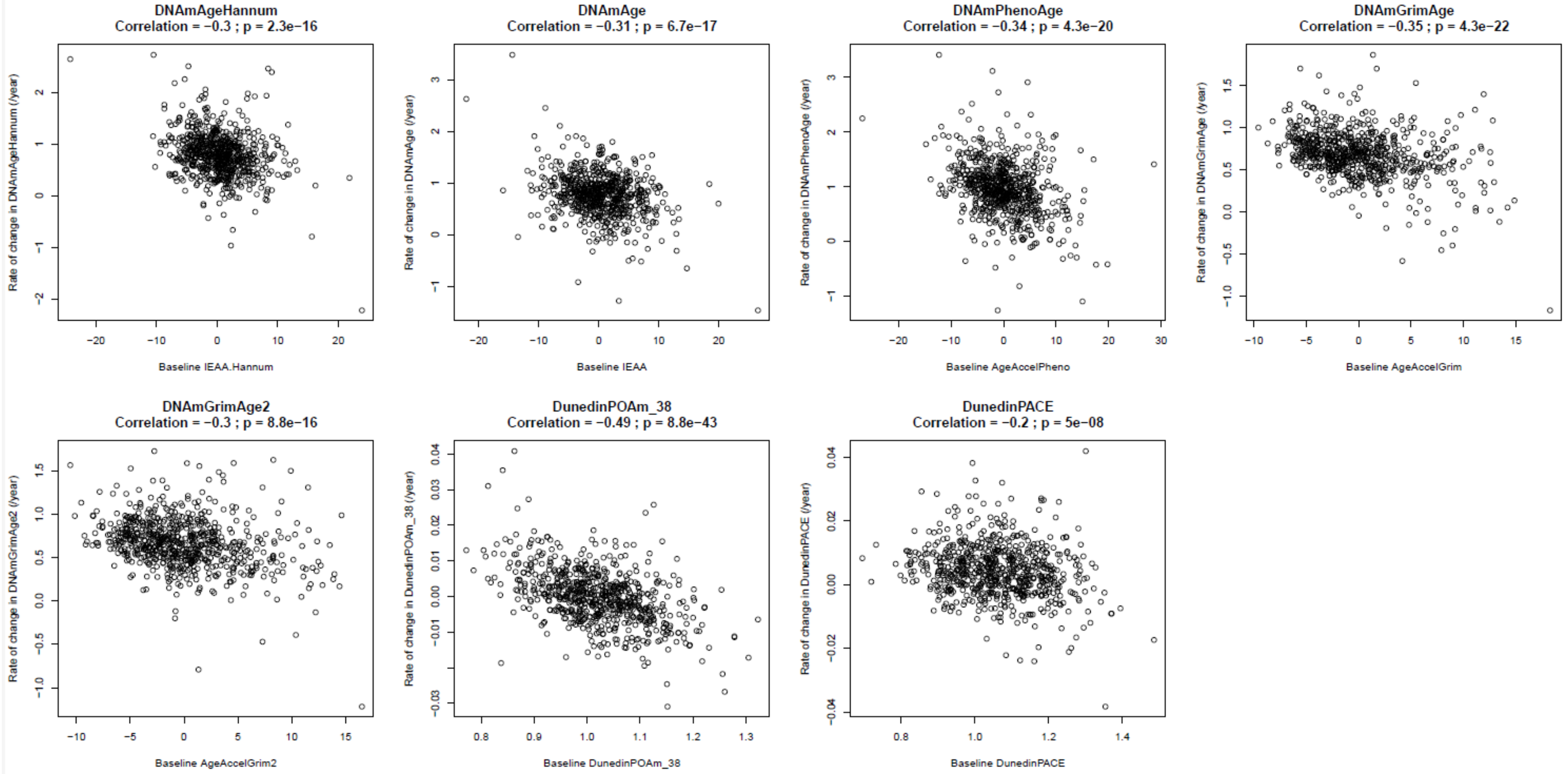


Because Hannum clock, Horvath clock, DNAmPhenoAge, DNAmGrimAge and DNAmGrimAge version 2 are highly correlated with chronological age, the chronological age adjusted variables are used for these measurements at the baseline. Specifically, IEAA.Hannum, IEAA, AgAccelPheno, AgeAccelGrim and AgeAccelGrim2 are used for Hannum clock, Horvath clock, DNAmGrimAge and DNAmGrimAge version 2 at the baseline.

**Title**: Longitudinal changes in epigenetic clocks predict survival in the InCHIANTI cohort

Supplemental Figure 2. Correlations between baseline epigenetic clocks and chronological age

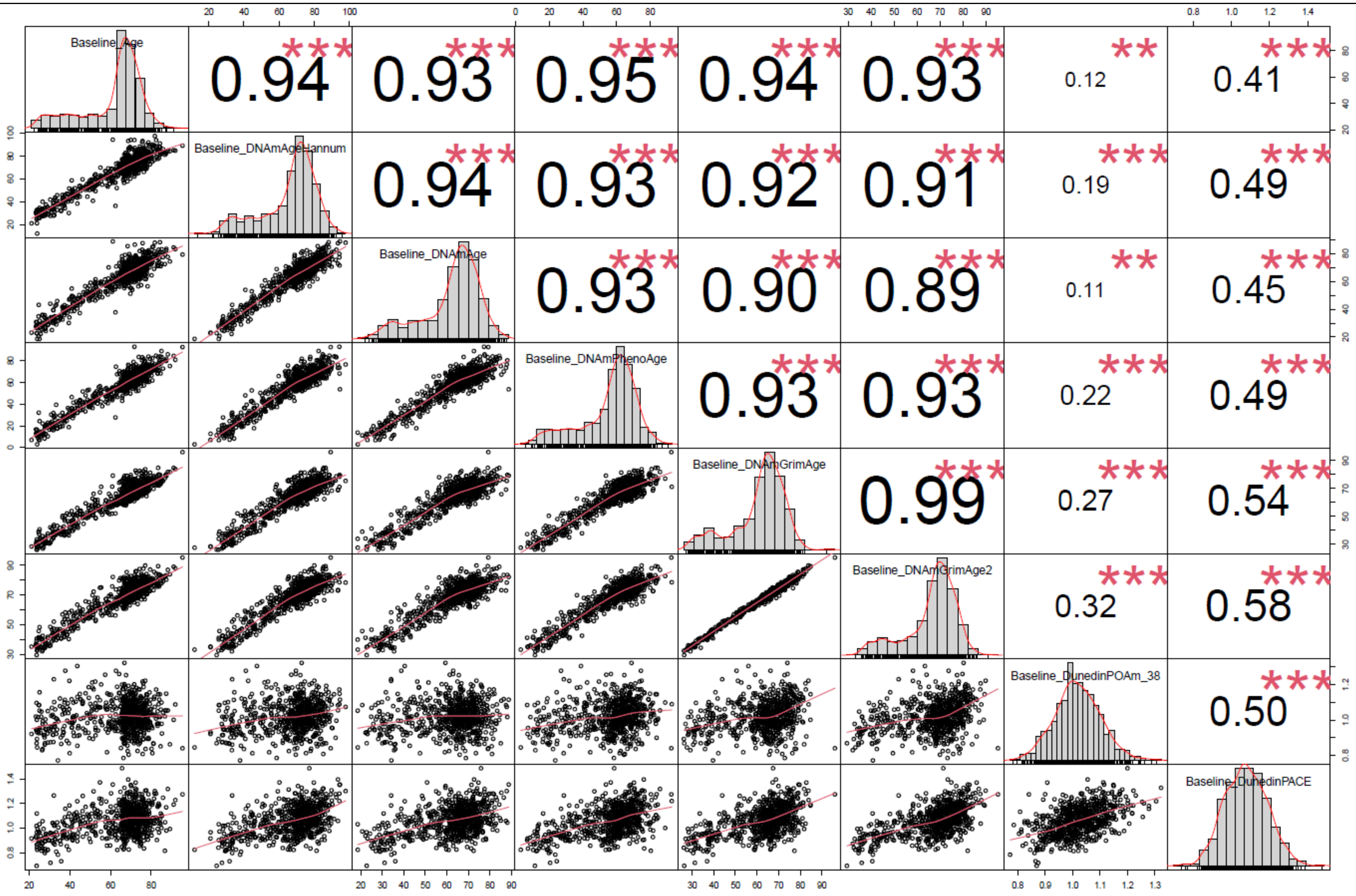

**Title**: Longitudinal changes in epigenetic clocks predict survival in the InCHIANTI cohort

This figure showed that Hannum clock, Horvath clock, DNAmPhenoAge, DNAmGrimAge and DNAmGrimAge version 2 were all highly correlated with chronological age. Thus, chronological age adjusted variables are used for these measurements at the baseline in this paper. Specifically, IEAA.Hannum, IEAA, AgAccelPheno, AgeAccelGrim and AgeAccelGrim2 are used for Hannum clock, Horvath clock, DNAmGrimAge and DNAmGrimAge version 2 at the baseline.

**Title**: Longitudinal changes in epigenetic clocks predict survival in the InCHIANTI cohort

**Title**: Longitudinal changes in epigenetic clocks predict survival in the InCHIANTI cohort

Supplemental Figure 3. Correlations between longitudinal changes in epigenetic clocks and baseline chronological age

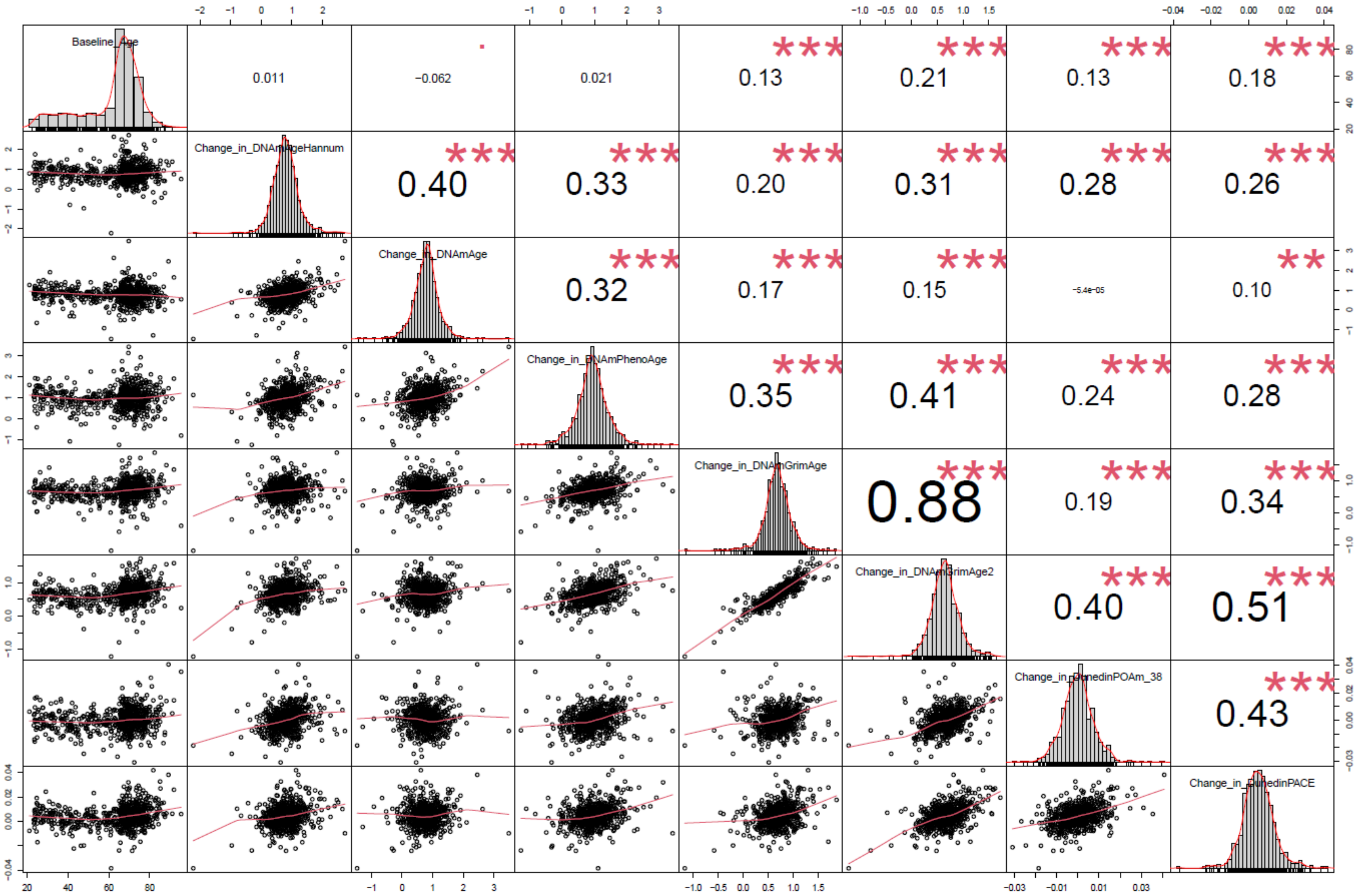

**Title**: Longitudinal changes in epigenetic clocks predict survival in the InCHIANTI cohort

**Title**: Longitudinal changes in epigenetic clocks predict survival in the InCHIANTI cohort

Supplemental Figure 4. Forest Plot for the Adjusted Hazard Ratios of Mortality when both Baseline and Longitudinal Changes are included in the model with additional adjustment for Life Simple Seven

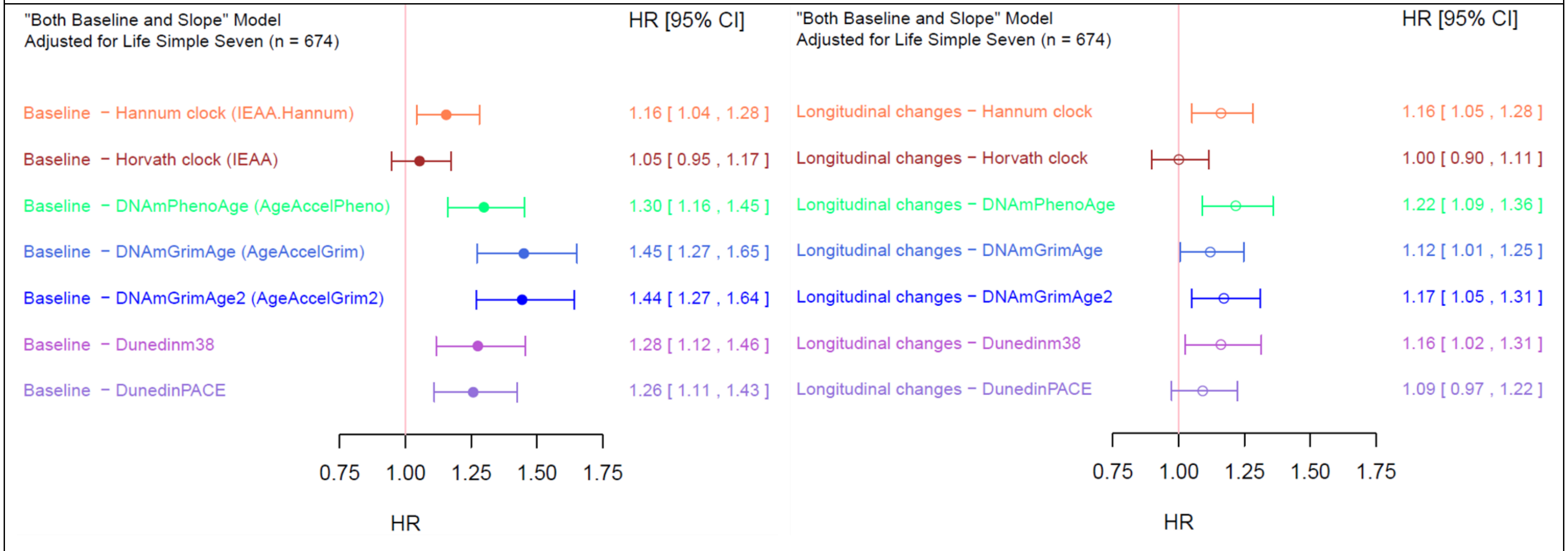


For Supplemental Figure 4, the hazard ratios are derived from the model including baseline epigenetic markers of aging, annual rate of longitudinal changes in epigenetic markers of aging, chronological age, sex, study site, and Life Simple Seven. Because Hannum clock, Horvath clock, DNAmPhenoAge and DNAmGrimAge are highly correlated with chronological age, the chronological age adjusted variables are used for these measurements at the baseline. Specifically, IEAA.Hannum, IEAA, AgAccelPheno, AgeAccelGrim and AgeAccelGrim2 are used for Hannum clock, Horvath clock, DNAmPhenoAge, DNAmGrimAge and DNAmGrimAge version 2 at the baseline in the model, respectively. To facilitate the interpretation and comparison, all the coefficients are standardized.

**Title**: Longitudinal changes in epigenetic clocks predict survival in the InCHIANTI cohort

Supplemental Figure 5. Forest Plot for the Adjusted Hazard Ratios of Mortality of PC clocks when both Baseline and Longitudinal Changes are included in the model

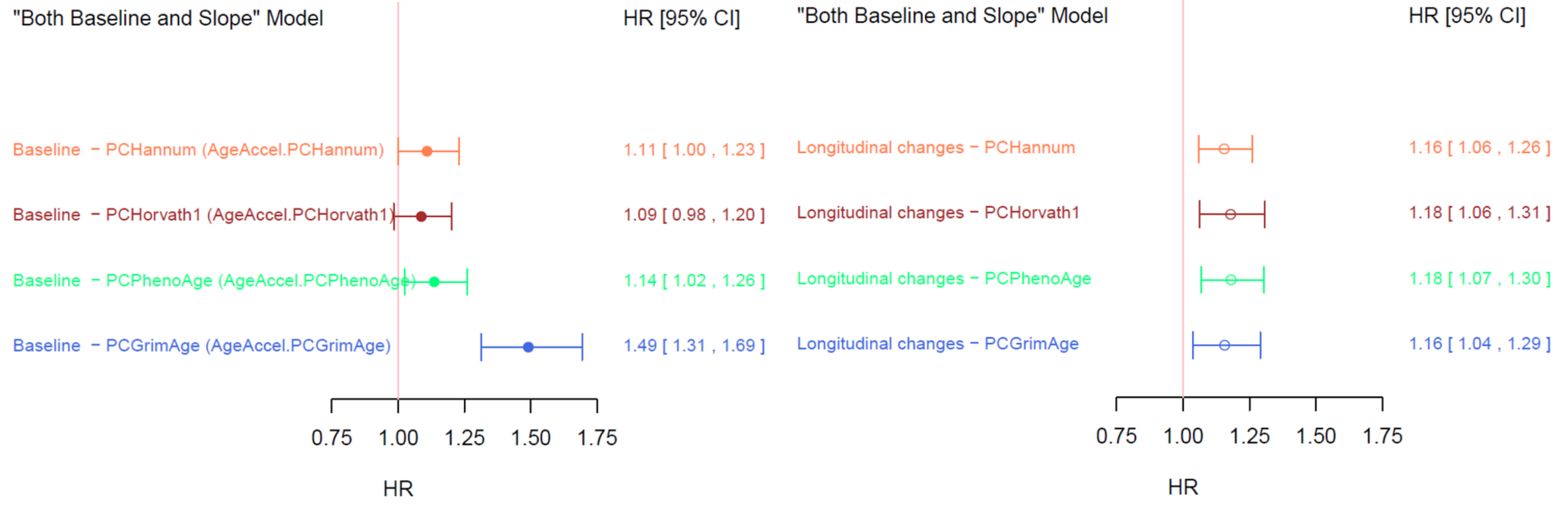


For Supplemental Figure 5, the hazard ratios are derived from the model including baseline age acceleration of PC clock, annual rate of longitudinal changes in PC clock, chronological age, sex, study site. Because these PC clocks are highly correlated with chronological age, the chronological age adjusted variables are used for these measurements at the baseline. Specifically, AgeAccel.PCHannum, AgeAccel.PCHorvath1, AgeAccel.PCPhenoAge, AgeAccel.PCGrimAge are used for PCHannum, PCHorvath1, PCPhenoAge, PCGrimAge at the baseline in the model, respectively. To facilitate the interpretation and comparison, all the coefficients are standardized.
